# No decline in functional status or cognitive function during the first year after treatment initiation for early-stage breast cancer in women aged 75 years and older: a prospective observational matched cohort study

**DOI:** 10.64898/2026.10.14.26365509

**Authors:** W.G. van der Plas-Krijgsman, J.W. Blom, M.G.M. Derks, T. van Dalen, L. van Gerven, O.R. Guicherit, T.E. Lans, E.M.H. Linthorst-Niers, J.W.S. Merkus, A.J.E. Vulink, S.P. Mooijaart, N.A. de Glas, J. Gussekloo, G.J. Liefers, J.E.A. Portielje

## Abstract

**Background:** This study compared functional and cognitive trajectories in older women treated for early breast cancer with matched women without cancer.

**Methods:** Data were derived from two Dutch cohorts: the CLIMB study (women aged ≥75 years with operable breast cancer) and the ISCOPE study (community-dwelling women aged ≥75 years without cancer). Breast cancer patients were matched 1:2 to controls on age and baseline functional or cognitive status. Functional status was assessed using the Groningen Activity Restriction Scale (GARS), and cognitive function using the Mini-Mental State Examination (MMSE). Longitudinal changes over 12 months were analyzed using both unadjusted and confounder-adjusted linear mixed models.

**Results:** 168 breast cancer patients and 336 matched breast cancer-free controls were included with a mean age of 80 years. Most breast cancer patients had stage 0-I or II disease and received surgery and/or radiotherapy; 48% received adjuvant endocrine therapy, chemotherapy was rare (<1%). At baseline, both cohorts were comparable in age and functional or cognitive status, although there was a higher comorbidity burden in the control cohort. No significant differences in functional or cognitive decline were observed between the cohorts over 12 months.

**Conclusions:** In older women, early breast cancer and its treatment did not lead to accelerated decline in functional status or cognitive function within the first year compared to matched older women without cancer. These findings suggest that, for relatively fit older patients, breast cancer treatment may not accelerate functional decline in the short term.

## 1. Introduction

Due to the rapid aging of the population, an increasing proportion of individuals are reaching advanced age. Older adults often experience gradual decline in physical and cognitive functioning, which may compromise their independence and quality of life. This functional decline can be influenced by various factors, including comorbidity, polypharmacy, reduced physical activity, and depressive symptoms, as shown in previous population-based studies.^1, 2^ However, the rate and extent of this decline vary considerably between individuals.

In line with the aging population, breast cancer is predominantly a disease of older women, with incidence rising strongly after the age of 65.^3^ In older women diagnosed with breast cancer, the abovementioned age-related changes may interact with the effects of cancer and its treatment, further increasing vulnerability. Additional declines in physical performance and cognition have been observed following chemotherapy and endocrine therapy.^4, 5^ Previous studies have also demonstrated increased risks of treatment-related toxicity, hospitalization, and functional decline in older patients compared to their younger counterparts^6, 7^ Yet, it remains unclear to what extent these changes reflect normal aging versus treatment-induced effects.

In order to determine the best choice of treatment for the individual older patient, understanding the potential influence of different cancer treatments on functional status and cognitive function is crucial. These insights may help to differentiate between treatment-related effects and expected decline due to natural aging, and can guide patient selection and optimize personalized treatment approaches to improve outcomes and preserve functional status and quality of life.

The aim of this study was therefore to compare the trajectories of functional status and cognitive function in women aged 75 years and older treated for early-stage breast cancer during the first year following treatment initiation, compared to (age and baseline functional or cognitive status) matched older women without breast cancer.

## 2. Methods

### 2.1 Design and study population

#### Climb study cohort

Breast cancer patients were selected from the Climb Every Mountain (CLIMB) study, a multicenter prospective longitudinal cohort study of women aged 70 years and older with primary operable breast cancer (stage 0-III) who were recruited between 2013 and 2018 from nine Dutch hospitals (N=637). Details of the CLIMB study have been published elsewhere.^8, 9^ Exclusion criteria were a previous breast cancer diagnosis, stage IV disease, advanced dementia or inability to read Dutch.

At baseline, all patients underwent a comprehensive geriatric assessment as part of standard care before start of primary treatment, with data collected on age, comorbidities (registered according to the Charlson Comorbidity Index (CCI))^10^, medication use, and various geriatric domains including functional status and cognitive function. Functional status was assessed using the Groningen Activity Restriction Scale (GARS)^11^, which consists of eleven items on ADL and seven items on IADL, resulting in a total score between 18 and 72, with higher scores indicating worse functional status. Cognitive function was assessed using the Mini-Mental State Examination (MMSE).^12^ Baseline tumor characteristics and data on primary treatment were retrieved from the medical charts. After start of primary treatment, follow-up visits took place at 3, 6, 12 and 24 months, consisting of multiple geriatric assessments, including the GARS questionnaire and MMSE. For the present analysis 12 month measurements were used.

#### ISCOPE study cohort

Older women without breast cancer were selected from the Integrated Systemic Care for Older People (ISCOPE) study, a large health-care innovation trial conducted in the South-West region of the Netherlands. Details of this study have been published elsewhere.^2, 13^ In brief, all persons aged 75 years and older, derived from 59 general practices between September 2009 and September 2010, were asked to participate (N=11,476). Exclusion criteria were people with terminal illness or an expected life expectancy of ≤3 months. A total of 7,285 participants completed the ISCOPE screening questionnaire, consisting of 23 questions on four different health domains (somatic, functional, psychological and social).^2^ Based on the screening tool outcome, a selection of participants was visited at home by a research nurse for additional data collection at baseline and at 12-month follow-up, including data on comorbidity and medication use, and assessment of functional status (using GARS) and overall cognitive function (using MMSE). The selection comprised 15% of all participants with problems in 0 or 1 domain, 60% with problems in 2 domains, and all participants with problems in 3 or 4 domains were selected.

### 2.2 Procedures and data

#### Inclusion criteria for this comparative study

For this study, we included participants from both cohorts who were female, 75 years and older and who completed the baseline geriatric assessment and at least one follow-up assessment in the next 12 months For patients from the ISCOPE cohort, patients who had a malignancy at baseline were excluded.

#### Matching

Breast cancer patients from the CLIMB cohort were automatically matched to the breast cancer-free participants from the ISCOPE cohort at a ratio of 1 case to 2 controls, with priority given to exact matches. We implemented matching with replacement, meaning that a single control could be matched to multiple treated CLIMB patients. This approach was chosen to minimize bias and ensure the closest possible matches. Matching was performed separately for the functional status and cognitive function analyses because these represent distinct domains of health with different baseline distributions. Simultaneous matching on both GARS and MMSE scores, in addition to age, would have substantially reduced the number of suitable matched controls and increased the likelihood of less precise or inconsistent matches, thereby compromising the efficiency and validity of the analyses. For the comparison of functional status over time, matching was based on age (with a range restriction of 2 years) and baseline GARS score (with a range restriction of 2 points). For the comparison of cognitive function over time, matching was based on age (range restriction of 2 years) and baseline MMSE score (range restriction of 2 points).

#### Comparison of baseline patient characteristics

The following baseline characteristics were available at baseline in both cohorts: age, living situation, marital status, education, and hospital admission within the last year. For the comparable geriatric measures, comorbidity was subdivided into 6 categories that were reported in both cohorts: diabetes, cardiovascular comorbidity (defined as heart failure or (prior) myocardial infarction or cerebrovascular accident), pulmonary disease (defined as asthma or chronic obstructive pulmonary disease), musculoskeletal comorbidity (defined as either presence of arthrosis or osteoporosis), psychiatric comorbidity (defined as depression or anxiety disorder) and sensory deficits (defined as any report of poor vision or cataract and any report of hearing problems or use of a hearing aid). These six categories formed the comorbidity count (subdivided into 4 groups: 0 comorbidities, 1-2, 3-4, and 5-6 comorbidities). Polypharmacy was defined as use of 4 or more daily medications.

For the CLIMB patients, tumor stage (categorized into stage 0-I, stage II and stage III), surgical treatment (either mastectomy, breast conserving surgery, or no surgery), and adjuvant radiotherapy, endocrine therapy and chemotherapy was reported. All baseline characteristics were obtained prior to surgery or prior to start of primary endocrine therapy in case patients did not receive surgical treatment.

#### Comparison of functional status and cognitive function

For the CLIMB patients, follow-up data on functional status was available at 3, 6, and 12 months after primary treatment initiation, and follow-up data for cognition was available at 6 and 12 months. For the matched ISCOPE participants, follow-up data for both outcomes was available at 12 months. For functional status, the GARS scores from both cohorts were compared and categorized into three groups (<19 points: independent; 19-28 points: partially dependent; and ≥29 points: fully dependent). If one answer was missing, the average of the other answers was calculated and given for the missing question. If more than 1 answer was missing, the GARS score was defined as unknown. For cognitive functioning, the MMSE scores that were collected in both cohorts were compared and categorized into three groups (≥28 points: normal mental status; 24 – 27 points: partially impaired mental status; and <24 points: impaired mental status). In case of missing answers, if less than 10% of the total score was missing, the maximum score was given for the missing question. If ≥10% was missing, the MMSE score was defined as unknown.

#### Outcomes

The main outcomes of this study were functional status (according to GARS) and cognitive function (according to MMSE) after 12 months in older patients with primary treatment for breast cancer and in matched controls.

### 2.3 Statistical analysis

All statistical analyses and matching were performed in SPSS version 25. Descriptive characteristics were reported using frequencies and proportions (%) for categorical variables and means with standard deviation (sd) for age. Pearson chi-squared test was used to assess differences in baseline characteristics between the cohorts. A p-value of less than 0.05 was considered statistically significant. Functional status and cognition were analyzed by composing graphs of the mean outcomes per follow-up visit with corresponding 95% confidence interval (CI) for patients with and without breast cancer. Linear mixed models (LMMs) were used to compare longitudinal changes in functional status and cognition between both cohort over time. Use of LMMs ensures that incomplete cases are included in the analysis, assuming that the data is missing at random.^14^ Functional status and cognitive function were separately analyzed as dependent variables, with study cohort (patient = CLIMB or control = ISCOPE), time (follow-up at 3, 6, and 12 months), and their interaction (time*cohort) as fixed effects. Models were adjusted for predefined confounders, based on clinical knowledge and previous research.^15^ For functional status, the predefined confounders were: education, baseline MMSE score, polypharmacy, comorbidity count, musculoskeletal comorbidity and psychiatric comorbidity. For cognitive functioning, the predefined confounders were: education, baseline GARS score, polypharmacy, comorbidity count, cardiovascular comorbidity, psychiatric comorbidity and vision and sensory deficits. We estimated the beta-coefficients with 95% CI and p-values.

## 3. Results

In total, we included 168 older breast cancer patients aged 75 years and older from the CLIMB cohort and 336 age-matched controls from the ISCOPE cohort. Tumor and treatment characteristics of the included CLIMB patients are presented in **Figure 1**. Most patients had either stage 0-I (46%) or stage II (41%) breast cancer. 46% of all patients underwent a mastectomy as primary treatment, 45% had breast conserving surgery, and 15% received no surgical treatment. Almost half of the patients received adjuvant radiotherapy (49%) and endocrine treatment (48%), and only 1% was treated with adjuvant chemotherapy.

**Figure 1.**
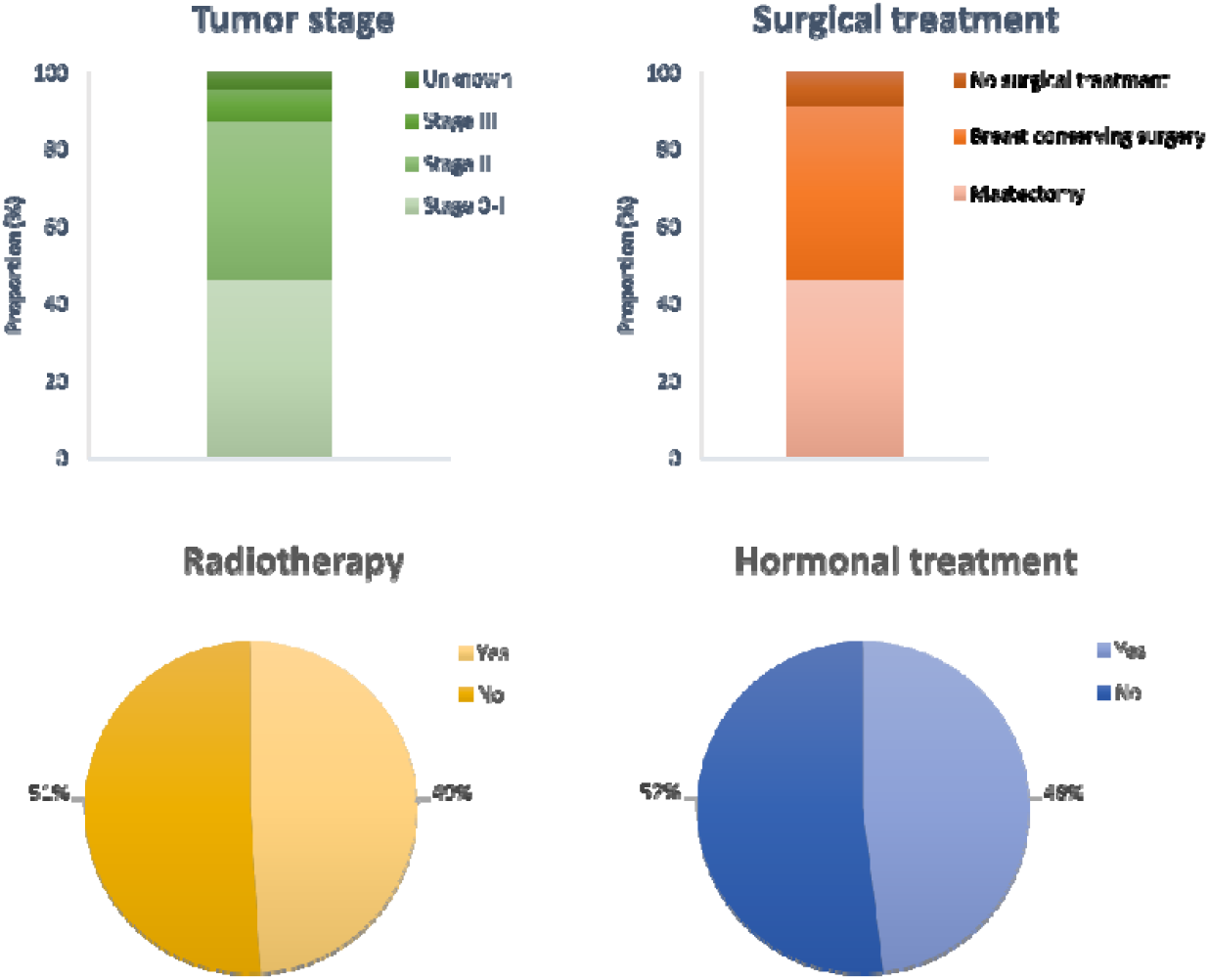
Tumor and treatment characteristics of the included CLIMB patients.

### Functional status over time

For the assessment of functional status over time, baseline characteristics of the CLIMB patients (N=164) and their matched ISCOPE comparisons (N=322) are presented in **Table 1**. Mean age was 80 years (SD 4) in both cohorts, with similar baseline GARS scores (p=0.944). The majority of women lived independently (>93% in both cohorts). More women in the ISCOPE control cohort were widowed (56% versus 43%, p=0.013), less educated (low education in 38% versus 21%, p<0.001), and had better baseline MMSE scores (normal mental status in 75% versus 67%, p=0.016). Polypharmacy was reported in more than half of the women in both cohorts. Comorbidities were more present in the ISCOPE control cohort (p<0.001 for comorbidity count), with significantly more cardiovascular and musculoskeletal comorbidity (both p<0.001).

**Table 1.** Baseline characteristics of Climb participants and their ISCOPE matched comparisons by age and baseline functional status (GARS)

|  | Climb<br>(N=164) |  | ISCOPE<br>(N=328) |  | *p-value* |
| --- | --- | --- | --- | --- | --- |
| Age (years) |  |  |  |  |  |
| Mean (sd) | 80.3 | 4.3 | 80.3 | 4.2 | 0.955 |
| Age in categories | n | % | n | % |  |
| 75-79 | 75 | 45.7 | 144 | 43.9 |  |
| 80-84 | 57 | 34.8 | 120 | 36.6 |  |
| 85-89 | 26 | 15.9 | 54 | 16.5 |  |
| 90+ | 6 | 3.7 | 10 | 3.0 | 0.954 |
| Baseline GARS score (ADL/IADL) |  |  |  |  |  |
| Independent <19 | 53 | 32.3 | 111 | 33.8 |  |
| Partially dependent 19 t/m 28 | 79 | 48.2 | 154 | 47.0 |  |
| Fully dependent ≥29 | 32 | 19.5 | 63 | 19.2 | 0.944 |
| Living situation* |  |  |  |  |  |
| Independent | 152 | 93.3 | 315 | 96.0 |  |
| Assisted living | 11 | 6.7 | 13 | 4.0 | 0.178 |
| Marital status * |  |  |  |  |  |
| Married or living together | 62 | 40.8 | 112 | 34.1 |  |
| Widow | 66 | 43.3 | 185 | 56.4 |  |
| Divorced/unmarried/other | 24 | 15.8 | 31 | 9.5 | 0.013 |
| Education**<sup>a</sup> |  |  |  |  |  |
| Low | 33 | 21.4 | 124 | 37.8 |  |
| Medium | 84 | 54.5 | 155 | 47.2 |  |
| High | 37 | 24.0 | 49 | 14.9 | <0.001 |
| Unplanned hospital admission in past year* |  |  |  |  |  |
| No | 128 | 82.6 | 286 | 87.2 |  |
| Yes | 27 | 17.4 | 42 | 12.8 | 0.176 |
| **Baseline MMSE score\*** |  |  |  |  |  |
| Normal mental status $\geq 28$ | 105 | 66.9 | 245 | 75.4 | |
| Partially impaired mental status 24 t/m 27 | 43 | 27.4 | 75 | 23.1 |  |
| Impaired mental status $< 24$ | 9 | 5.7 | 5 | 1.5 | 0.016 |
| **Polypharmacy\*** |  |  |  |  |  |
| < 4 medications | 66 | 42.6 | 153 | 46.6 |  |
| $\geq 4$ medications | 89 | 57.4 | 175 | 53.4 | 0.402 |
| **Diabetes** |  |  |  |  |  |
| No | 130 | 79.3 | 279 | 85.1 |  |
| Yes | 34 | 20.7 | 49 | 14.9 | 0.106 |
| **Cardiovascular comorbidity<sup>b</sup>** |  |  |  |  |  |
| No | 125 | 76.2 | 202 | 61.6 |  |
| Yes | 39 | 23.8 | 126 | 38.4 | <0.001 |
| **Pulmonary disease<sup>c</sup>** |  |  |  |  |  |
| No | 147 | 89.6 | 279 | 85.1 |  |
| Yes | 17 | 10.4 | 49 | 14.9 | 0.161 |
| **Musculoskeletal comorbidity<sup>d</sup>** |  |  |  |  |  |
| No | 115 | 70.1 | 155 | 47.3 |  |
| Yes | 49 | 29.9 | 173 | 52.7 | <0.001 |
| **Psychiatric comorbidity<sup>e</sup>** |  |  |  |  |  |
| No | 151 | 92.1 | 299 | 91.2 |  |
| Yes | 13 | 7.9 | 29 | 8.8 | 0.732 |
| **Sensory deficits<sup>f</sup>** |  |  |  |  |  |
| No | 90 | 54.9 | 160 | 48.8 |  |
| Yes | 74 | 45.1 | 168 | 51.2 | 0.202 |
| **Comorbidity count** |  |  |  |  |  |
| 0 | 46 | 28.0 | 42 | 12.8 |  |
| 1 or 2 | 88 | 53.7 | 205 | 62.5 |  |
| 3 or more | 30 | 18.3 | 81 | 24.7 | <0.001 |

\*results presented without unknowns
<sup>a</sup>educational level: low defined as primary school or less; medium defined as secondary school or middle vocational education; high defined as higher vocational education or university
<sup>b</sup>cardiovascular comorbidity: defined as prior myocardial infarction or cerebrovascular accident (CVA) or heart failure
<sup>c</sup>pulmonary disease: defined as either presence of asthma or chronic obstructive pulmonary disease (COPD)
<sup>d</sup>musculoskeletal comorbidity: defined as either presence of arthrosis or osteoporosis
<sup>e</sup>psychiatric comorbidity: defined as either (prior) depression or anxiety disorder
<sup>f</sup>sensory deficits: defined as any report of poor vision or cataract and any report of hearing problems or use of a hearing aid

In the CLIMB cohort, 4 patients were excluded due to missing GARS scores at baseline. Follow-up at 12 months was completed in 110 patients (67%) in the CLIMB cohort and in all the ISCOPE controls. Linear mixed models showed no statistical significant difference in functional status between the CLIMB breast cancer patients and the ISCOPE control cohort (unadjusted: b=0.68; 95% CI -0.75 - 2.12; p=0.349, and adjusted: b=1.38; 95% CI -0.09 - 2.84; p=0.07).) The longitudinal trajectories over time did show a small, but statistically significant decrease in mean GARS score in the CLIMB patient cohort, indicating slight improvement of functional status during follow-up (unadjusted p=0.004 and adjusted p=0.02) (**Fig 2A**).

**Figure 2.**
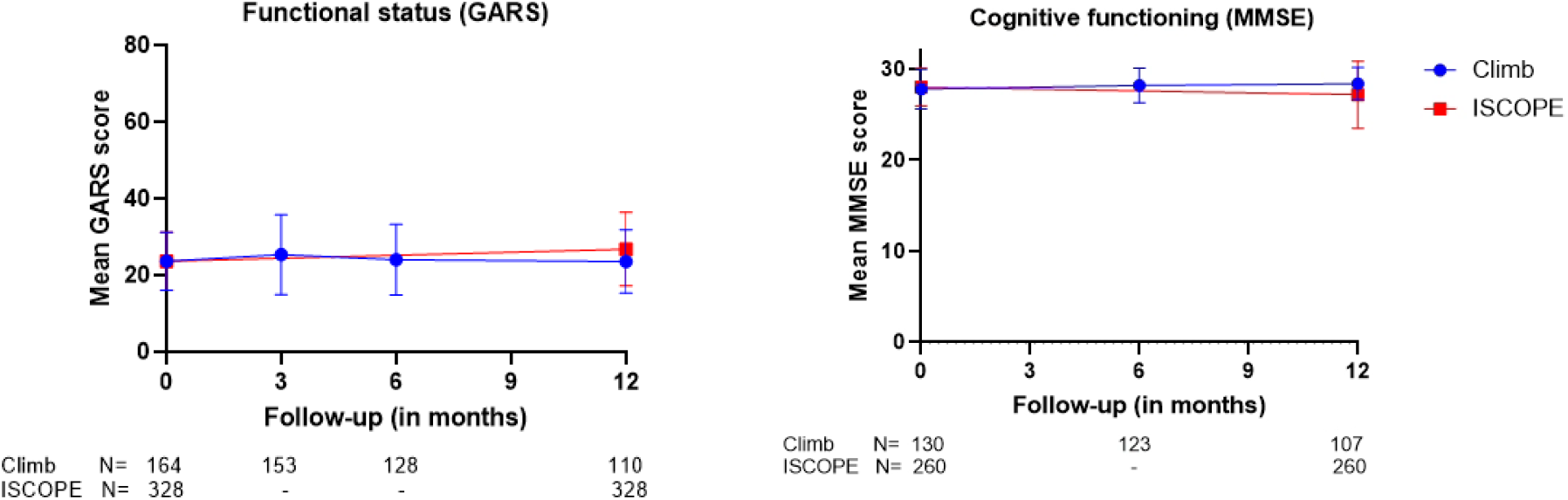
Longitudinal changes in functional status (A) and cognitive function (B)

### Cognitive function over time

For the assessment of cognitive function over time, baseline characteristics are presented in **Table 2**. Mean age was 80 years (SD 4), with similar baseline MMSE scores (p=0.917). Most women lived independently (>91% in both cohorts). In the ISCOPE control cohort more women were widowed (47% versus 40%, p=0.05), less educated (low education in 38% versus 19%, p<0.001), and had better functional status at baseline according to mean GARS scores (fully dependent in 49% vs 16%, p<0.001). In the ISCOPE cohort, there was more polypharmacy (54% in CLIMB vs 66% in ISCOPE, p=0.023), and more comorbidities (p<0.001 for comorbidity count), with significantly more cardiovascular comorbidity (p<0.001), pulmonary disease (p=0.012), musculoskeletal comorbidity (p<0.001), and psychiatric comorbidity (p=0.032).

**Table 2.** Baseline characteristics of Climb participants and their ISCOPE matched comparisons by age and baseline cognitive functioning (MMSE)

|  | Climb<br>(N=130) |  | ISCOPE<br>(N=260) |  | *p-value* |
| --- | --- | --- | --- | --- | --- |
| **Age (years)** |  |  |  |  |  |
| Mean (sd) | 79.9 | 4.2 | 79.9 | 4.1 | 0.963 |
| **Age in categories** | **n** | **%** | **N** | **%** |  |
| 75-79 | 64 | 49.2 | 121 | 46.5 |  |
| 80-84 | 47 | 36.2 | 94 | 36.2 |  |
| 85-89 | 15 | 11.5 | 37 | 14.2 |  |
| 90+ | 4 | 3.1 | 8 | 3.1 | 0.896 |
| **Baseline MMSE score** |  |  |  |  |  |
| Normal mental status ≥28 | 90 | 69.2 | 180 | 69.2 |  |
| Partially impaired mental status 24 t/m 27 | 35 | 26.9 | 72 | 27.7 |  |
| Impaired mental status <24 | 5 | 3.8 | 8 | 3.1 | 0.917 |
| **Living situation\*** |  |  |  |  |  |
| Independent | 123 | 95.3 | 237 | 91.2 |  |
| Assisted living | 6 | 4.7 | 22 | 8.5 |  |
| Nursing home | 0 | 0.0 | 1 | 0.4 | 0.301 |
| **Marital status \*** |  |  |  |  |  |
| Married or living together | 46 | 40.4 | 100 | 38.5 |  |
| Widow | 45 | 39.5 | 123 | 47.3 |  |
| Divorced/unmarried/other | 23 | 20.2 | 37 | 14.2 | 0.05 |
| **Education<sup>a</sup>** |  |  |  |  |  |
| Low | 22 | 19.1 | 99 | 38.1 |  |
| Medium | 65 | 56.5 | 119 | 45.8 |  |
| High | 28 | 24.4 | 42 | 16.1 | <0.001 |
| **Hospital admission in past year\*** |  |  |  |  |  |
| No | 97 | 83.6 | 204 | 78.5 |  |
| Yes | 19 | 16.4 | 56 | 21.5 | 0.242 |
| **Baseline GARS score (ADL/IADL)\*** |  |  |  |  |  |
| Independent <19 | 45 | 34.9 | 32 | 12.3 |  |
| Partially dependent 19 t/m 28 | 64 | 49.6 | 100 | 38.5 |  |
| Fully dependent ≥29 | 20 | 15.5 | 128 | 49.2 | <0.001 |
| **Polypharmacy\*** |  |  |  |  |  |
| < 4 medications | 57 | 46.3 | 89 | 34.2 |  |
| ≥4 medications | 66 | 53.7 | 171 | 65.8 | 0.023 |
| **Diabetes** |  |  |  |  |  |
| No | 102 | 78.5 | 207 | 79.6 |  |
| Yes | 28 | 21.5 | 53 | 20.4 | 0.791 |
| **Cardiovascular comorbidity<sup>b</sup>** |  |  |  |  |  |
| No | 104 | 80.0 | 144 | 55.4 |  |
| Yes | 26 | 20.0 | 116 | 44.6 | <0.001 |
| **Pulmonary disease<sup>c</sup>** |  |  |  |  |  |
| No | 119 | 91.5 | 213 | 81.9 |  |
| Yes | 11 | 8.5 | 47 | 18.1 | 0.012 |
| **Musculoskeletal comorbidity<sup>d</sup>** |  |  |  |  |  |
| No | 89 | 68.5 | 94 | 36.2 |  |
| Yes | 41 | 31.5 | 166 | 63.8 | <0.001 |
| **Psychiatric comorbidity<sup>e</sup>** |  |  |  |  |  |
| No | 120 | 92.3 | 220 | 84.6 |  |
| Yes | 10 | 8.7 | 40 | 15.4 | 0.032 |

| Sensory deficits<sup>f</sup> |  |  |  |  |  |
| --- | --- | --- | --- | --- | --- |
| No | 71 | 54.6 | 124 | 47.7 |  |
| Yes | 59 | 45.4 | 136 | 52.3 | 0.197 |
| Number of comorbidities |  |  |  |  |  |
| 0 | 38 | 29.2 | 34 | 13.1 |  |
| 1 or 2 | 70 | 53.8 | 122 | 46.9 |  |
| 3 or more | 22 | 17.0 | 104 | 30.0 | <0.001 |
| *\*results presented without unknowns* |  |  |  |  |  |
| *<sup>a</sup>educational level: low defined as primary school or less; medium defined as secondary school or middle vocational education; high defined as higher vocational education or university* |  |  |  |  |  |
| *<sup>b</sup>cardiovascular comorbidity: defined as prior myocardial infarction or cerebrovascular accident (CVA) or heart failure* |  |  |  |  |  |
| *<sup>c</sup>pulmonary disease: defined as either presence of asthma or chronic obstructive pulmonary disease (COPD)* |  |  |  |  |  |
| *<sup>d</sup>musculoskeletal comorbidity: defined as either presence of arthrosis or osteoporosis* |  |  |  |  |  |
| *<sup>e</sup>psychiatric comorbidity: defined as either (prior) depression or anxiety disorder* |  |  |  |  |  |
| *<sup>f</sup>sensory deficits: defined as any report of poor vision or cataract and any report of hearing problems or use of a hearing aid* |  |  |  |  |  |

In the CLIMB cohort, 38 patients were excluded due to missing MMSE scores at baseline (N=7) or missing follow-up data (N=31). Follow-up at 12 months was completed in 107 patients (82%) in the CLIMB cohort and in all ISCOPE controls. Linear mixed models showed similar mean MMSE scores in both cohorts (unadjusted: b=0.02; 95% CI -0.55 – 0.50; p=0.931, and adjusted: b=-0,42; 95% CI -0.99 – 0.16; p=0.16) (**Fig 2B**). Over time, the CLIMB cohort showed a small but statistically significant increase in mean MMSE score during follow-up (unadjusted p=0.003 and adjusted p=0.005).

## 4. Discussion

Our results showed no significant differences in functional or cognitive decline in the first year after treatment initiation between patients with breast cancer and matched breast cancer-free controls. Together with previous research, these findings provide a more detailed perspective on functional outcomes in older patients.

Previous research has identified the vulnerability of older patients with breast cancer for both functional and cognitive decline following treatment. A study by Derks et al. illustrated a persistent decline in physical functioning in the years after endocrine treatment, unlike younger patients who showed stabilization of recovery.^4^ Physical functioning was assessed according to Metabolic Equivalent of Task (MET) hours, which might partially explain the differences with our findings, together with a longer follow-up period of 2 years. Another study by Lemij et al, showed a small decline in physical activity and a small increase in ADL/IADL dependency (measured with GARS) in the first 5 years after breast cancer diagnosis, with more prominent functional decline after the first year.^16^

Concurrent with physical decline, cognitive impairment is frequently reported in older patients after breast cancer treatment. Lange et al. reported that up to 50% of older patients with early-stage breast cancer showed objective cognitive decline following adjuvant therapy, with particularly individuals treated with chemotherapy at risk.^5^ Older breast cancer survivors with cognitive impairment after treatment reported greater frailty, reduced physical function, and lower quality of life.^17^

Previous studies have demonstrated an association between cancer treatments and increased risk of frailty and reduced quality of life.^18-20^ However, as some authors have pointed out, factors such as age, multimorbidity, and psychosocial variables may independently influence these declines, making it difficult to attribute them fully to cancer treatment.^21^ Furthermore, multiple studies have shown that physical and cognitive decline are not always strongly correlated with breast cancer diagnosis or its treatment. Instead, geriatric characteristics including advanced age, comorbidities, low educational level, depression and impaired mobility have been independently associated with both physical and cognitive deterioration in this patient population.^22-24^ These findings are consistent with prior research examining functional decline and disability in the general older population.^1, 2^

Our findings suggest that, in a relatively fit older population in which chemotherapy was rarely administered and approximately half of the patients received endocrine therapy, clinically meaningful cognitive and functional decline may be less prevalent. The treatment pattern in this study reflects current clinical practice in the Netherlands, where few older women with hormone-receptor positive breast cancer receive chemotherapy.^25, 26^ Our results are consistent with similar studies reporting that endocrine therapy alone was not associated with substantial functional of cognitive decline in the first years following treatment.^22, 23^ Furthermore, results from the recently published ASTER 70s trial, a large phase III randomized trial by Brain et al, demonstrated that although the addition of adjuvant chemotherapy to endocrine treatment did not improve survival outcomes in older breast cancer patients, there was a substantial increase in adverse events and decline in health-related quality of life.^27^ Functional deterioration and adverse outcomes in older patients therefor differ considerably when chemotherapy is part of the treatment regimen.^5, 28^

### Strengths and Limitations

A major strength of this study is the prospective longitudinal design and the use of matched comparisons from two large cohorts, in order to differentiate between the effects of cancer treatment and aging-related effects on functioning. Validated measures of functional (GARS) and cognitive (MMSE) functioning were applied uniformly in both cohorts.

However, several limitations are evident. The follow-up data for the CLIMB cohort was incomplete for a proportion of patients. Since we included only patients whose data on functional status and cognitive function was available at baseline and at least one follow-up visit during the first 12 months, this may have introduced selection bias. The follow-up duration of 12 months may be insufficient to identify late-onset and progressive functional or cognitive decline. The relatively short follow-up time was determined by the study design, as the control cohort only had available follow-up data up to one year. In previously published research derived from the ISCOPE cohort similar limitations have been described due to the short follow-up, and specifically a high rate of drop-outs mostly due to functional decline that could not be included in the study due to lack of follow-up data at 12 months.^2^ However, other CLIMB studies without matched controls have reported longer-term outcomes of functional and cognitive function, providing valuable insights beyond this timeframe.^22-24^

Patient selection may have contributed to our findings. The CLIMB cohort consisted predominantly of older women that were physically fit enough to undergo breast cancer surgery and additional systemic treatment. This could indicate that more frail older patients may have been excluded for treatment or were less likely to participate in the CLIMB study, potentially leading to more favorable outcomes, as baseline frailty and physical fitness are important predictors of treatment tolerance and recovery in older breast cancer patients.^15^ On the contrary, participants in the ISCOPE study may have been more frail at baseline, with lower socio-economic status and a higher comorbidity count. The method of matching used in this study helps to address these baseline differences between the cohorts, but it cannot completely rule out remaining differences or account for unmeasured variables.

Lastly, only a very small number of patients received chemotherapy, which may limit how well the results apply to patients receiving more aggressive treatment. Yet, as mentioned earlier, we know that these percentages are similar for Dutch clinical practice, where chemotherapy treatment for early breast cancer stages in women aged 75 years and older is substantially lower compared to other countries in Europe.^25, 26^

### Clinical Implications and Future Research

Our findings provide insight that in relatively fit older women with early breast cancer treated with surgery, radiotherapy and hormonal therapies, significant declines in functional status and cognitive function during the first year need not be anticipated. Future research should focus on longer-term follow-up to investigate prolonged effects of breast cancer treatments on functional outcomes, as well as inclusion of more frail older patients, to increase the generalizability of outcomes. Frail patients often experience higher rates of treatment-related complications and greater declines in functional status and cognitive function after therapy, underscoring the need for comprehensive geriatric assessment to guide treatment decisions.^29^

## 5. Conclusion

In this study, older women with early-stage breast cancer primarily treated with surgery, radiotherapy and adjuvant endocrine treatment, did not experience greater functional or cognitive decline over one year compared to matched controls without cancer. These findings suggest that, for relatively fit older patients, current breast cancer treatment approaches according to Dutch clinical practice may not accelerate functional and cognitive decline in the short term. Longer follow-up studies are needed to further understand the long-term impact of breast cancer and different treatment modalities on functional outcomes.

## Data availability statement

The data used for this study is available upon reasonable request.

## Ethical approval

For both the CLIMB study and ISCOPE trial, written informed consent was obtained from all participants. The CLIMB study was approved by the medical ethics committee of the Leiden University Medical Center (LUMC) (CCMO:NL43463.058.13). The ISCOPE trial was approved by the LUMC medical ethics committee, and is registered in the Netherlands Trial Register (NTR1946).

## Declaration of generative AI and AI-assisted technologies

During the preparation of this work the author(s) used ChatGPT in order to improve language and readability of the text. ChatGPT was also used to refine the graphical abstract. After using this tool, the author(s) reviewed and edited the content as needed and take(s) full responsibility for the content of the published article.

